# Noninvasive optical monitoring of cerebral blood flow and EEG Spectral responses after Severe Traumatic Brain Injury: A Case Report

**DOI:** 10.1101/2021.07.14.21260436

**Authors:** Chien-Sing Poon, Benjamin Rinehart, Dharminder S. Langri, Timothy M. Rambo, Aaron J. Miller, Brandon Foreman, Ulas Sunar

## Abstract

Survivors of severe brain injury may require care in a neurointensive care unit (neuro-ICU), where the brain is vulnerable to secondary brain injury. Thus, there is a need for noninvasive, bedside, continuous cerebral blood flow monitoring approaches in the neuro-ICU. Our goal is to address this need through combined measurements of EEG and functional optical spectroscopy (EEG-Optical) instrumentation and analysis to provide a complementary fusion of data about brain activity and function. The present case demonstrates in a patient with traumatic brain injury, noninvasive cerebral blood flow transients can be recorded that correlate with gold-standard invasive measurements and with the frequency content changes in the EEG data during clinical care.

## 1. INTRODUCTION

Traumatic brain injury (TBI) is a significant public health burden, contributing to 30% of all injury-related deaths in the U.S. and resulting in long-term disability for more than 3 million Americans at an annual cost of $80 billion [1]. There is a significant interest in developing effective management strategies to improve clinical outcomes. Survivors of severe TBI often require critical care, during which the brain is vulnerable to secondary injuries including anoxia, ischemia, and edema that result in part from an uncoupling of cerebral blood flow (CBF) from metabolic demand. A major goal of neurocritical care is to monitor the brain to detect and minimize these secondary brain injuries. There is a need for real-time, noninvasive, multimodal measurements for intervention guidance.

Several clinical technologies for imaging are available, including computed tomography (CT) and magnetic resonance imaging (MRI). While these are noninvasive, imaging provides only single snapshots in time, and are not suited for continuous, long-term monitoring at the bedside neuro-ICU settings [2]. In contrast, continuous monitoring in most ICUs is limited to continuous assessments of cardiopulmonary function. Invasive probes to measure intracranial pressure (ICP) are recommended for select patients according to the Brain Trauma Foundation Guidelines [1,3]. We recently described a standardized method for multimodality monitoring (MMM) which includes not only ICP but a thermal diffusion probe (Hemedex, Inc; Cambridge, MA) to measure CBF measurements from brain tissue directly[4]. However, these methods require drilling a burr hole through the skull and placement of probes directly into cortex, conferring a risk for hemorrhage while sampling only one area of the frontal lobe. Laser Doppler flowmetry (LDF) measures CBF from a very small volume of tissue, and LDF devices have not been FDA-approved for clinical use in humans [5]. Transcranial Doppler ultrasound (TCD) measures the velocity of flow within large cerebral arteries, but cannot directly measure CBF, is limited by skull thickness, and is challenging to use for longitudinal monitoring due to the need for stable ultrasonic probe orientation [6].

Continuous EEG (cEEG) is capable of detecting seizures, periodic discharges, and spreading depolarizations leading to secondary brain ischemia and may contain relevant prognostic information in patients after moderate or severe TBI [7–10]. Currently, cEEG monitoring is recommended by the international neurocritical care community for use in patients following TBI. It has been recognized that characteristic changes occur during EEG in response to brain ischemia in correlation with CBF and oxygen metabolism [11,12]. When CBF becomes compromised, the metabolic and electrical activity of cortical neurons is impacted leading to alternations in the frequency content of the EEG [12]. However, the use of EEG for ischemia monitoring is not widespread despite commercially available quantitative software. Thus, there exists an unfilled niche for continuous, bed-side monitoring of cerebral blood flow within the vulnerable cortex in the neuro-ICU setting.

A diffuse optical technique, diffuse correlation spectroscopy (DCS), is ideal as a monitor of CBF in high-risk populations, such as those with TBI [13–15]. DCS has been used to provide noninvasive, continuous measurements of CBF in animals and in humans [13,15–17] and recently its use in neuro-ICU settings [6,18–20] has demonstrated that DCS measurements agree with clinically-established modalities. Further, we recently demonstrated the potential to use DCS to elucidate resting state functional connectivity (RSFC) [21,22]. Here we report continuous cerebral tissue blood flow measurements in a TBI patient by using optical diffuse correlation spectroscopy. Clinically-standard electroencephalogram (EEG) auxiliary recordings provided a complementary fusion of data about brain activity and function.

## 2. MATERIALS AND METHODS

### 2.1 Study Design and Patient Details

This is a case report from a patient with severe TBI admitted to the Neuroscience Intensive Care Unit (NSICU) in an American College of Surgeons (ACS) level I trauma center. Invasive multimodality neuromonitoring devices were placed per our clinical standard and time-locked with cEEG and systemic physiologic data as previously described. All care was provided according to national (ACS) guidelines. Written informed consent was provided by a legally authorized representative prior to study procedures as approved by the Institutional Review Board at the University of Cincinnati.

A man that is in his early forties with no significant past medical history was admitted after being struck by a motor vehicle. Upon arrival of Emergency Medical Services, he was noted to have extensor posturing and was intubated in the field. In the Emergency Department, his Glasgow Coma Scale (GCS) score was 3T and he had bilaterally nonreactive pupils. Noncontrast head CT demonstrated an 8 mm left subdural hematoma with 1.2 cm left-to-right midline shift (**Figure 1**). Hypertonic saline was given, resulting in the return of bilateral pupillary reactivity and improvement in clinical exam to 6T. He was taken emergently to the operating room where he underwent left decompressive hemicraniectomy and clot evacuation. In addition, he was found to have several grade-1 blunt cerebrovascular injuries, cervical spinal hyperextension injury without cord signal change, and multiple orthopedic fractures. On post-trauma day 1 and post-operative day 0, he underwent placement of multimodality intracranial monitoring.

**FIGURE 1.**
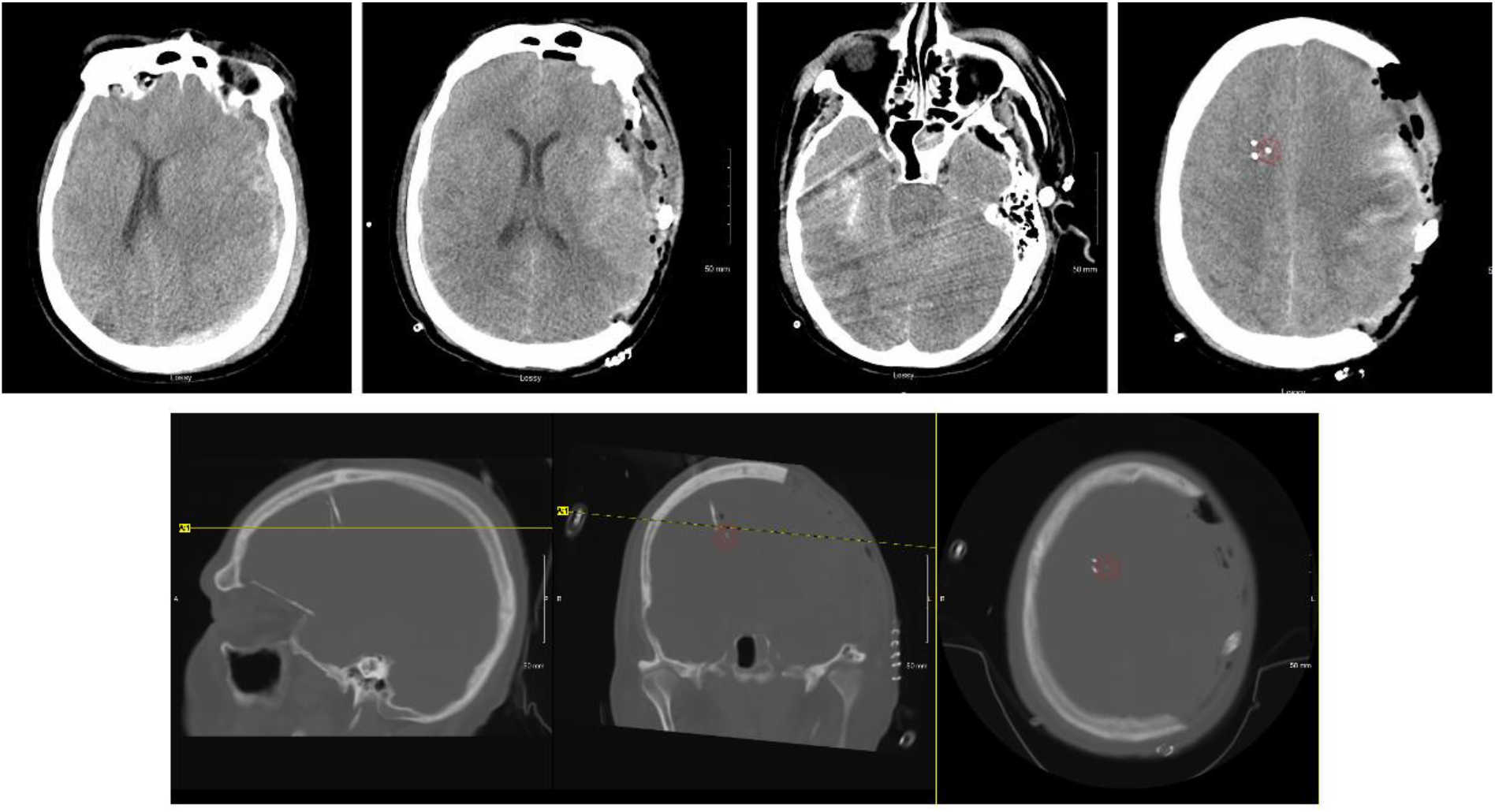
Computed Tomography (CT) head imaging. Top row (left to right): A) Pre-operative imaging at presentation demonstrated left subdural hematoma with midline shift. B-D) Post-operative imaging after decompressive hemicraniectomy and evacuation of the acute hemorrhage demonstrating peri-hematoma contusion but resolution of the midline shift. An area of right temporal contusion is also seen with relative compression of the cisterns surrounding the brainstem. Intracranial monitoring devices were placed in the contralateral hemisphere approximately 2.5cm under the inner table of the skull within subcortical white matter. The location of the Bowman Perfusion probe is highlighted by a red circle. Bottom row (left to right): Sagittal, coronal, and axial sections of the post-operative CT imaging windowed to highlight dense structures including the intracranial monitoring devices. The Bowman Perfusion probe is highlighted with a red circle on coronal and axial sections.

### 2.2 Optical Technique

Optical data were acquired using a custom-built DCS system, as we published previously [21,22]. In summary, the custom-made DCS instrument employed a continuous-wave laser source (785 nm CrystaLaser; Reno, NV, USA) with a coherence length longer than 10 m, eight NIR-optimized superconducting Nanowire single-photon counting detectors (QuantumOpus, LLC; Novi MI, USA), and an 8-channel auto-correlator board (Correlator.com). A multi-mode fiber (1000 um core diameter) was used to guide the 785 nm laser light to the scalp and few-mode fiber (8.2 mm core diameter) coupled quantum nanowire single-photon counting modules collected the light. Photodetector outputs are fed into a correlator board that computed light intensity temporal autocorrelation functions that are recorded by a computer using a custom LabVIEW software. We used four DCS detectors at 2.7 cm, three detectors at 2.5 cm to average the signal, and one detector at shortest separation of 8mm to obtain the scalp signal. The custom optical probe was based on a foam pad that incorporated prisms, enabling 90-degree light delivery and detection for optimal probe-tissue contact. DCS data were acquired at 1 Hz.

For the quantification of the CBF using DCS technique, we assumed an optical absorption parameter of 0.15 cm^-1^ and a scattering parameter of 10 cm^-1^. The normalized diffuse electric field temporal autocorrelation function (g_1_ (r,τ)) was extracted from measured normalized intensity temporal autocorrelation function (g_2_ (r,τ)) and then it is fitted to an analytical solution of the diffusion equation to estimate the blood flow index (BFi) parameter.

All clinical monitoring data were exported into common format (Matlab) for analysis using CNS Envision (Version 1.00.00) with the CNS Data Conversion plugin (Moberg Solutions, Inc; Ambler, PA). From scalp EEG recordings, we chose the electrode channel most closely overlying the region studied by the optical probe (F4). We incorporated EEG data from the nearby depth electrode, chosing the single channel recording of the highest amplitude and best quality. To get the density spectral array (DSA), we used Welch’s method to generate power spectra with 8 segments and 50% overlap from 1-20 Hz. Each segment was windowed with a Hamming window and over 4s epochs for scalp and depth channels. The power of the theta frequency band (4-8 Hz) was obtained by applying a 1-min moving average filter. The alpha-delta ratio (ADR) [23] was calculated by taking the ratio of the power of the alpha frequency band to the delta frequency band with a 2-min moving average filter.

## 3. RESULTS

Clinical multimodality monitoring initially demonstrated decompressed physiology with resolving cerebral edema. Regional cerebral blood flow (rCBF) was highly correlated with mean arterial blood pressure (MAP), suggesting impaired regulation, although improvements were seen in the absolute rCBF from post-trauma day 1 to 2. Cerebral microdialysis demonstrated mitochondrial dysfunction with the development of relatively low interstitial brain glucose concentrations between days 2 and 3. Brain tissue oxygen monitoring did not respond to increased fractional inspiration of oxygen (FiO2), and therefore PbtO2 values were deemed not accurate measurements of dissolved oxygen tension (PtO2) within the monitored region of tissue. Confirmatory head CT demonstrated adequate probe placement without peri-catheter hematoma (**Figure 1**).

On post trauma day 2, intravenous administration of labetalol 10 mg resulted in an abrupt decreases in mean arterial blood pressure (MAP; 98 mmHg to 61 mmHg); cerebral perfusion pressure (CPP; 86 mmHg to 51 mmHg); ICP (13 mmHg to 10 mmHg); and heart rate (HR; 105 to 91 bpm) (**Figure 2**).

**FIGURE 2.**
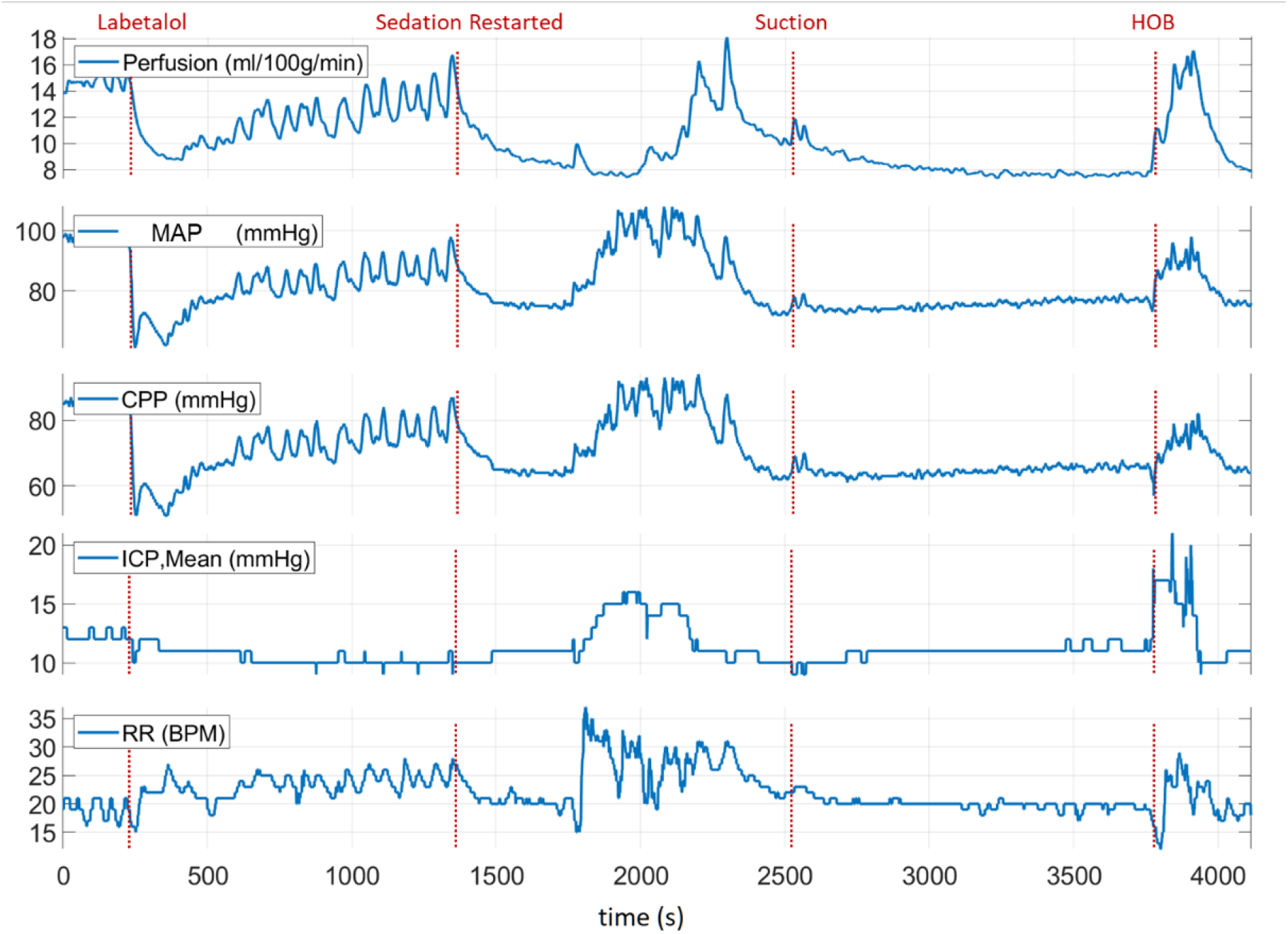
Clinical multimodality monitoring of the TBI patient on post trauma day 2. The red dashed line indicates clinically-relevant events annotated during the care of the patient.

Noninvasive optical probe was placed close to scalp EEG channel F4 to monitor the CBF (**Figure 3a**). During the optical spectroscopy recording, the optical (DCS rCBF) measurements indicated a similar trend as compared to the rCBF measured with the invasive TDF probe. After administration of labetalol, there was a sharp decrease in both rCBF (42.1%, ∼15.2 ml/100g/min t ∼8.7 ml/100g/min) and DCS rCBF (63.6%, ∼1.1 cm^2^/s to ∼0.4 cm^2^/s) (**Figure 3b**). Sedative medications, propofol at 40 mcg/kg/min and fentanyl 200 mcg/hr, were paused for a period of approximately 20 minutes as blood pressure recovered, resulting in vasomotor oscillations in rCBF, MAP, and HR signals as observed in **Figure 2**. Subsequently, the sedation was restarted where a decrease in rCBF (∼48.8%, ∼16.8 ml/100g/min to ∼8.5 ml/100g/min) with a time-locked decrease in DCS rCBF (cm^2^/s) (∼45.4%, ∼0.99 cm^2^/s to ∼0.54 cm^2^/s) was observed (**Figure 3b**). In the middle of the recording period, an airway suctioning procedure was performed, resulting in a brief increase in rCBF (20.2%, ∼9.9 ml/100g/min to ∼11.9 ml/100g/min) and DCS rCBF (17.3%, ∼0.75 cm^2^/s to ∼0.88 cm^2^/s) (**Figure 3b**). Toward the end of the recording period, the head of the bed (HOB) was lowered for patient repositioning, resulting in an increase in MAP and ICP, and a doubling of the rCBF (115%, ∼7.9 ml/100g/min to ∼17.0 ml/100g/min) (**Figure 2**). The DCS rCBF simultaneously increased (66%, ∼0.59 cm^2^/s to ∼0.98 cm^2^/s).

**FIGURE 3.**
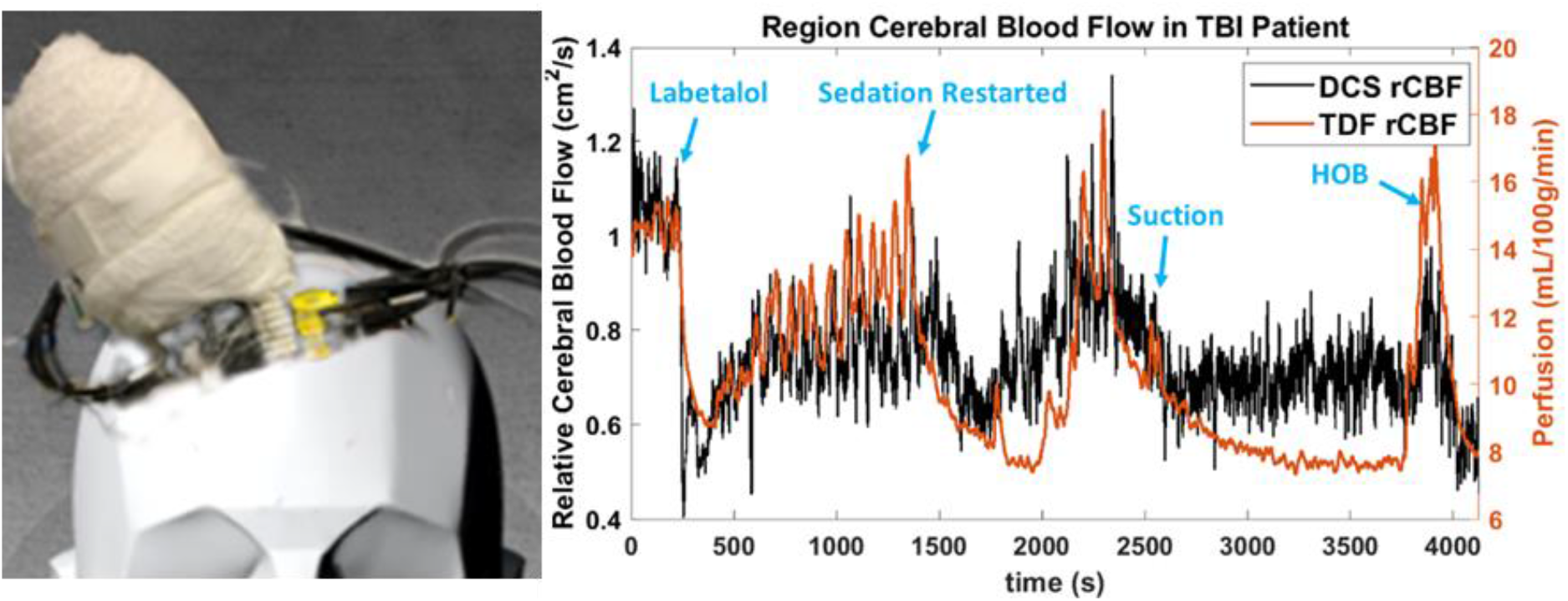
The feasibility of noninvasive optical blood flow measurements after severe traumatic brain injury (sTBI). (**a**) Invasive probe, scalp EEG, and noninvasive optical probe placement; (**b**) Measured blood flow with invasive, thermal-diffusion flowmetry (TDF rCBF) and noninvasive optical imaging (DCS rCBF). Sedation medication led to a substantial blood flow decrease, while HOB manipulation led to a significant blood flow increase, which could be measured by both techniques.

Next, we examined the EEG and ECoG responses to changes in rCBF, as summarized in **Figure 4** and **Figure 5**. Quantitative EEG signal analysis indicated reduction of higher frequencies and a decrease in ADR (%) and DSA (Hz), which aligned well with both clinical CBF and DCS derived CBFi. These findings suggest compromised cerebral perfusion pressure (CPP) (**Figure 5**).

**FIGURE 4.**
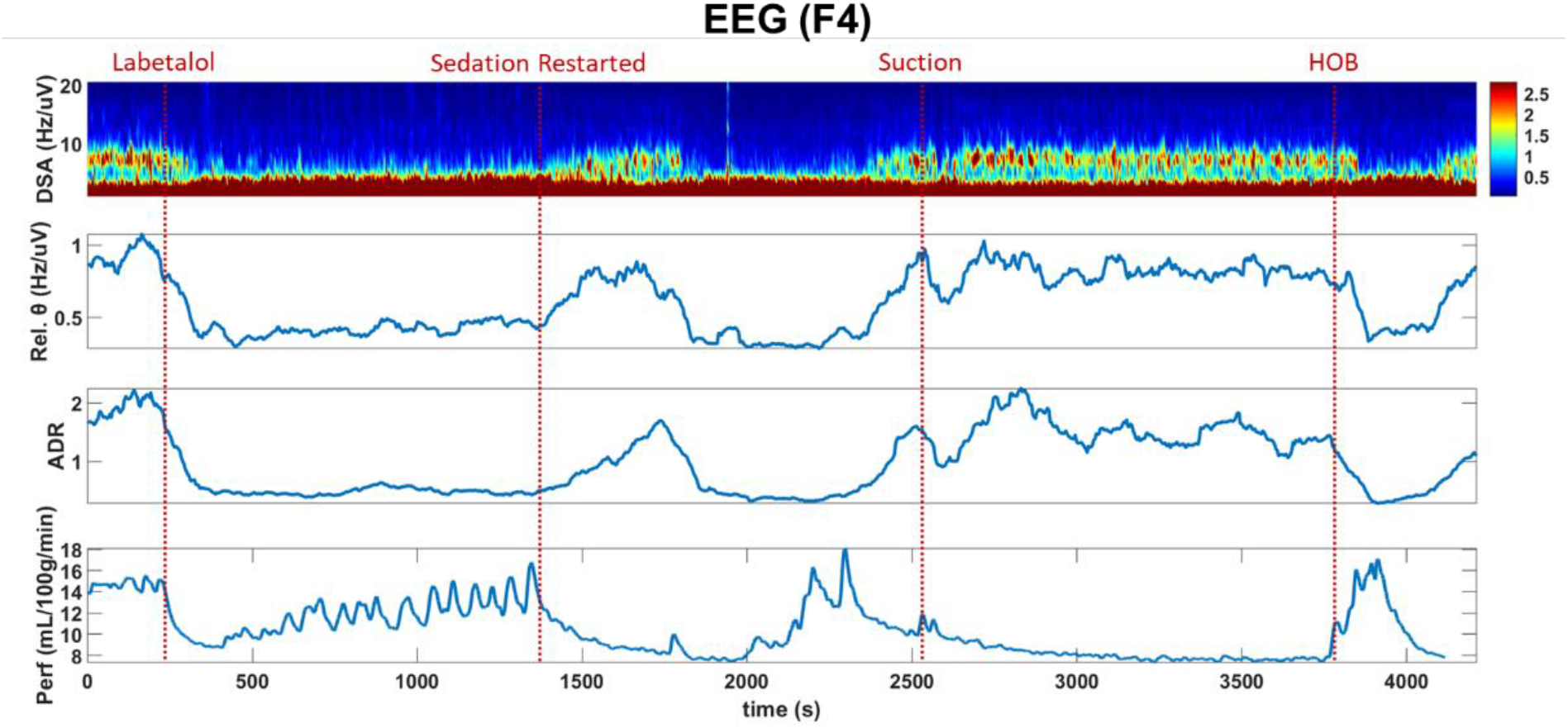
Scalp EEG (F4) recording correlates with rCBF. Fast Fourier transform of scalp EEG recordings along with rCBF (Perf). After labetalol administration, the rCBF decreases along with the high frequency content within the scalp EEG signal, as has been described during ischemia. In contrast, there is increasing high frequency activity after sedation is restarted despite a decrease in rCBF as might be expected with sedation, such as propofol, which lowers the metabolic demand of the tissue and is associated with diffuse higher frequency activity on EEG. This pattern is interrupted by suctioning and by manipulation of the head of the bed, with an inverse relationship between rCBF and higher frequency EEG activity as might be expected during arousal.

**FIGURE 5.**
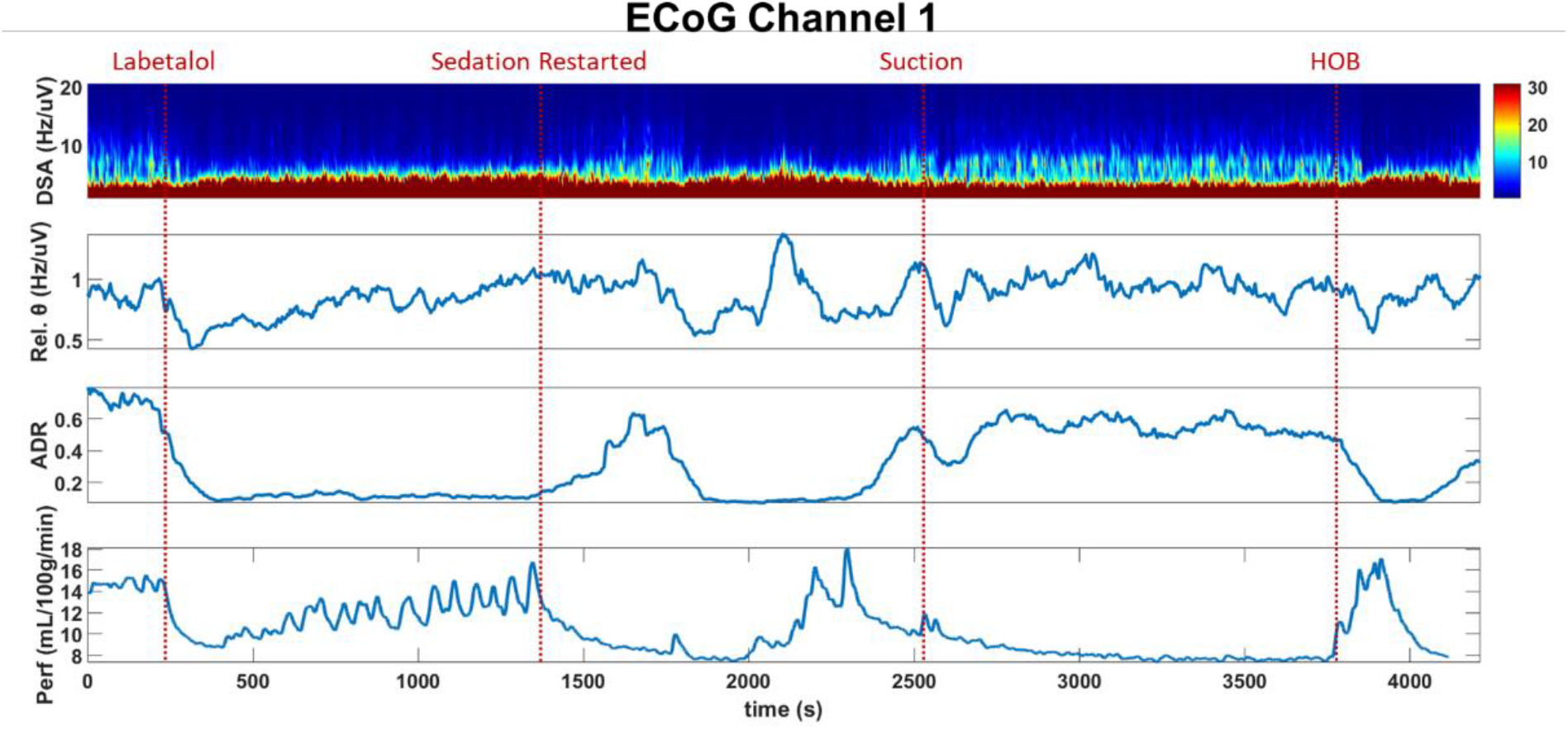
Depth EEG (ECoG) recording correlates with BF. Fast Fourier transform of depth electrode recordings along with rCBF (Perf). The relationship between regional blood flow and regional cortical post-synaptic potentials is similar to those observed on scalp EEG.

The patient subsequently required open reduction and fixation of the right femur and external fixation of both knees and anterior cervical decompression and fixation of the cervical spine. His hospital course was complicated by persistent respiratory failure with refractory hypoxia thought to be related in part to multi-segment rib fractures and pneumonia. He required tracheostomy and percutaneous gastrostomy prior to discharge to long-term acute care.

## 4. Discussion

We demonstrated here that optical blood flow measurements are capable of providing continuous, noninvasive monitoring of quantitative blood flow that can be coupled with EEG and other multimodality neuromonitoring parameters to provide important insights at the bedside. After sTBI, neurocritical care is aimed at limiting secondary injuries by closely monitoring the brain. Existing methods exhibit limit temporal or spatial sampling, limiting their utility. Optical imaging technologies have been promising but commercially available devices do not measure blood flow directly, leading to challenges in interpretation. We demonstrated that our method clearly correlated with invasive measurements of rCBF and further that our measurements informed the interpretation of EEG signals to diagnose a period of relative ischemia and the impact of arousal and sedation.

Invasive ICP monitoring (**Figure 5**) remains a cornerstone of neurocritical care. However, intracranial monitoring incurs a small but important risk for complications related to placement and devices are further limited by sampling from only a single region within the brain. Multiple modalities may be important to better understand the state of the monitored tissue relative to isolated measurements of ICP. As such, we previously published our clinical platform for providing multimodality monitoring as part of standard care, including measurements of rCBF using a thermal diffusion flowmetry device [4]. One of our main goals of this pilot study was to show the feasibility of providing continuous, longitudinal measurements of blood flow. In this study, we measured blood flow for approximately two hours, but our technique can continually measure CBF for 24 hours or more. A primary limitation was the need for space on the scalp which may be taken by numerous other devices, both invasive and noninvasive. However, optical fibers can be arranged close to EEG electrodes, as we showed here, and this can reduce the additional space requirements needed and allow for routine correlation of optical metrics with added spatial sampling in conjunction with EEG recordings.

Continuous EEG (cEEG) monitoring is currently recommended to detect electrographic seizures and may be useful for ischemia monitoring [12] based on characteristic changes that occur on EEG in response to decreases in blood flow oxygen metabolism [11,12]. As normal CBF declines, the EEG loses faster frequencies relative to slower frequencies as a crucial ischemic threshold is reached. Indeed, we observed that declines in the relative CBF index (**Figure 2**) were closely correlated with quantitative rCBF and both aligned with this expected EEG frequency response (**Figures 3 and 4**).

Importantly, we also highlighted that EEG frequency changes alone are non-specific – with decreases in faster frequency activity that occurred within the context of increased CBF during states of relative stimulation or arousal. Our noninvasive measurements of blood flow were adequate to allow for crucial contextualization of our observations from scalp and depth EEG.

Importantly, we highlighted the validity of our optical noninvasive measurements relative to direct, invasive thermal diffusion-based CBF measurements, which correlated well throughout the course of recording and during multiple clinical events. Optical measurements are performed by placing sensors on the scalp, which is a highly vascular tissue that can induce extracerebral signal contamination. Increasing the source detector separation to probe deeper within a tissue can occur at the cost of lower signal-to-noise (SNR) ratio. Superconducting nanowire detectors have much higher quantum efficiency (>90%) at ∼800nm compared to traditional single photon counting modules (e.g., Excelitas, ∼60%), which improved our SNR at longer source detector separations. Adding multiple fibers (multiple detectors) for each position to average the optical signal may further circumvent this issue but at an additional cost for each additional detector. We find that it is preferable that optical probes are adjusted for each patient individually in order to improve SNR at a minimal cost based on the monitoring required.

## 5. CONCLUSION

We described here a case study where optical imaging using DCS provided continuous, noninvasive blood flow changes which a) correlated well with direct, invasive measurements of rCBF, b) integrated with multimodality monitoring data including ICP and c) linked scalp and depth EEG with blood flow to highlight changes associated with ischemia, sedation, and arousal. These results support the clinical feasibility of our approach to noninvasive continuous bedside monitoring in the Neuro-ICU setting. In the future, this multimodal approach may provide neurological and physiological information to guide intervention to eliminate the risk of secondary brain injury.

## Data Availability

The data are stored locally, please contact the corresponding author to discuss.

## ACKNOWLEDGMENTS

We thank clinical nurses for their help during measurements and clinical interventions.

## AUTHOR CONTRIBUTIONS

U.S. and B.F. conceived the study. B.F., C.P., B.R., D.L. collected patient data. B.F. provided neuromonitoring expertise. U.S. and B.F. drafted the manuscript, and all authors contributed substantially to its revisions. U.S. and B.F. take responsibility for the paper as a whole.

## DATA AVAILABILITY STATEMENT

Data may be available via direct contacting via email request.

## INSTITUTIONAL REVIEW BOARD STATEMENT

The study was conducted according to the guidelines of the Declaration of Helsinki, and approved by the Institutional Review Board of University of Cincinnati.

## INFORMED CONSENT STATEMENT

Informed consent was obtained from all subject(s) involved in the study.

